# Comparative analysis of discriminative and generative natural language processing pipelines for automated prostate magnetic resonance imaging reports

**DOI:** 10.64898/2026.07.12.26357886

**Authors:** Daniel Lee, Nathan McCoy, Candace L. Haroldsen, Michael Gilkey, Sadna Verma, Anant Madabhushi, Saiju Pyarajian, Kara N. Maxwell, Nicholas G. Nickols, Matthew B. Rettig, Giuseppina Silvestri, Isla P. Garraway

## Abstract

**Objectives:** Natural language processing (NLP) can enable scalable extraction of clinically relevant information from unstructured radiology reports retrieved from electronic healthcare data warehouses, but reliance on externally hosted models may pose cost, privacy, and deployment challenges. We compared self-hosted discriminative and generative NLP pipelines for automated extraction of Prostate Imaging and Reporting Data System (PI-RADS) scores from multiparametric magnetic resonance imaging (mpMRI) reports used in prostate cancer risk assessment.

**Materials and Methods:** We identified 44,511 mpMRI reports across 68 Veterans Affairs (VA) healthcare systems. A stratified random sample of 1,973 reports was used to train, test, and evaluate multiple pipeline configurations combining Named Entity Recognition (NER) models and large language models (LLMs). Performance was assessed by accuracy of maximum PI-RADS extraction and processing speed using self-hosted implementations of spaCy NER, Transformers NER, and generative LLMs Llama 3, Qwen3, and Gemma3.

**Results:** Across the top 10 pipeline configurations, accuracy for maximum PI-RADS extraction ranged from 89.3% to 95.5%, with processing times spanning 150 milliseconds to 70 seconds per report. Generative LLM pipelines achieved the highest accuracy (up to 95.5%) but were substantially slower (2–70 seconds), whereas NER-based pipelines demonstrated lower accuracy (≈88.5%) with faster performance (50–150 milliseconds).

**Discussion:** Discriminative NER pipelines achieved high accuracy while offering advantages in speed and potential scalability. Accuracy gains from LLMs were accompanied by significantly higher computational cost, potentially limiting feasibility in high-volume clinical environments.

**Conclusion:** Discriminative methods were more efficient than generative models in annotating PI-RADS from mpMRI report text, providing insights into configurations for optimal clinical deployment when volume is a limiting factor. However, generative AI offered improved accuracy with less upfront development.

## Background and Significance

Multiparametric magnetic resonance imaging (mpMRI) of the prostate has become a fundamental component of prostate cancer detection and localization.^1-3^ The Prostate Imaging Reporting and Data System (PI-RADS) serves as the standardized assessment framework to identify regions of interest for targeted biopsies in mpMRIs.^4^ However, essential radiographic data generated from mpMRI exams, including PI-RADS scores, prostate volume, and PSA density determinations often exist in unstructured formats within radiology reports. Manual review and data abstraction of these report texts is a time-intensive process that could benefit from automated approaches. Recent advances in both discriminative and generative NLP models present opportunities to automate various aspects of prostate cancer screening and diagnosis workflows. Named Entity Recognition (NER) models excel at extracting specific clinical entities, while Large Language Models (LLMs) demonstrate capabilities in complex reasoning and text generation tasks. However, the optimal combination of these approaches for clinical applications for specific tasks in retrieving information, like PI-RADS from prostate mpMRI reports found in electronic health records, has not been previously evaluated comparatively.

The automated analysis of prostate mpMRI and pathology reports is emerging as a critical area bridging clinical decision support using machine learning methodologies.^5^ Recent work has focused on discriminative approaches to automate PI-RADS classification in radiology reports,^5,6^ as well as distinguish malignant from non-malignant tissue, and estimate Gleason grade in pathology reports.^7^ However, comparative analyses reveal inherent trade-offs and limitations, especially in balancing diagnostic accuracy with clinical interpretability. Several studies^8-11^ found that expert clinicians still outperform LLMs in diagnostic accuracy for cancer detection or classification, highlighting gaps in current automated approaches. However, the emergence of LLMs presents untapped potential for clinical applications of generative approaches for information extraction and inference. Existing literature lacks systematic comparisons between discriminative and generative paradigms for prostate cancer diagnosis workflows. Moreover, existing models have not been conducted on large, heterogeneous populations, which may undermine clinical utility in real-world settings and reinforce existing biases.^12^ While discriminative models excel in structured classification tasks, generative models offer advantages in contextual reasoning and multi-modal information synthesis-capabilities that remain underexplored in automated screening applications. Consequently, this gap in comparative analysis motivates our investigation into multiple text processing pipelines that leverage complementary strengths of both discriminative precision and generative contextual understanding for enhanced prostate cancer screening assessment. This study addresses the gap by systematically evaluating multiple text processing pipelines on a large-scale and in diverse populations. We present a comprehensive experimental framework that varies model configurations across critical pipeline components that can be run on standard hardware. Finally, one major consideration in this study was data privacy and computational accessibility. Some of the most performant models were run on a local laptop (Mac M3 with 48GB Ram), making the barrier for entry more affordable for low-income environments. It also alleviates risk related to privacy such as HIPAA by localizing computation to a single local computational resource.

## Materials and Methods

### Institutional Approval and Data Sources

Study approval was obtained through the VA Central Institutional Review Board (cIRB), which waived the informed consent requirement and authorization by the Health Insurance Portability and Accountability Act of 1996 because the study involved no patient contact and therefore minimal risk. We followed the Strengthening and Reporting of Observation Studies in Epidemiology (STROBE) reporting guideline.

This study included patients treated within the nationwide Veterans Affairs (VA) healthcare system from 2005 to 2024 who underwent prostate MRI (n= 81,904). Text from key sections of radiology reports (main report text and clinical impression text) were available in 44,511 documents retrieved from the VA Corporate Data Warehouse (CDW) accessed through the VA Multi-OMICS Analysis Platform for Prostate Cancer (VA-MAPP) database as previously described.^12,13^ Of the 44,511 documents, a random subset of 1,973, which were representative of variance in mpMRI report text creation and patient geographic variability, were selected for analysis. Radiology main report text and impression text were concatenated for unified processing and divided into 1,674 documents for training and testing named entity recognition systems with 299 documents held out for end-to-end pipeline evaluation.

### Experimental Design and Text Annotation

Our experimental framework allowed comparison of pipelines composed of variable discriminative NER models and/or generative LLMs components. NER models were trained on annotated data while the LLMs were applied without data annotation. Three independent annotators reviewed all documents to annotate the following entities: PI-RADS score (1-5), prostate dimensions, prostate volume, PSA, and PSA density (Supplementary Table 1).

For training the NER model, the main report and clinical impression texts from 1,674 of the 1,973 sample documents were split into training (80%, 1340 texts) and testing (20% 334 texts) sets. The testing set was used to validate the NER pipeline performance and ensure high quality predictions. End-to-end pipeline evaluation was performed using the remaining 299 documents held out for this purpose.

PI-RADS annotation for the end-to-end pipeline evaluation was performed by three annotators with high agreement (88% for both Cohen’s Kappa and Krippendorff’s alpha). The annotation label associated with each text was the maximum PI-RADS value explicitly written in the text. In cases where explicit PI-RADS value was not found, the text was annotated as PI-RADS “Unavailable” label (Supplementary Table 2).

The results obtained from the end-to-end pipelines were checked against the explicitly appearing maximum PI-RADS score in the text.

### Models and Pipeline Configurations

A variety of pipeline configurations and models were assessed. For each differing configuration and model, evaluation was performed against the 299 pipeline evaluation data set. All model pipeline configurations gathered the maximum PI-RADS value from an mpMRI text. Then, each text had a maximum PI-RADS score associated with the input, which is then checked against expected PI-RADS values that were annotated within the evaluation set.

## Results

Three pipeline configurations were evaluated to extract maximum PI-RADS using 299 mpMRI holdout texts (Figure 1). The maximum PI-RADS value is a risk-adverse clinical assessment that can affect patient care decisions (i.e., whether to undergo a diagnostic biopsy to detect prostate cancer). Therefore, automating and highlighting the value is important for streamlining diagnosis.

Two of the configurations (pipeline 1 & 2) employed a two-step procedure, where an information extraction phase was followed by a function that selects the maximum PI-RADS entity detected. In pipeline 1, all the entities supported by the NER model were extracted. A function to determine the maximum PI-RADS extracted from the PI-RADS Score entities is applied, and the output is formatted as a single numerical value. In pipeline 2, all the entities supported by a multi-shot prompt given to an LLM were extracted. A function to determine the maximum PI-RADS extracted from the PI-RADS Score entities was applied. The output was formatted as a single numerical value, discarding hallucinations or incorrect formatting. The generative LLM extraction supports all the same entity types as the discriminative NER. The third configuration (pipeline 3) utilized a single generative model with a multi-shot prompt to infer the maximum PI-RADS. The output was formatted as a single numerical value, discarding hallucinations or incorrect formatting. Each pipeline extracted the maximum PI-RADS value and was tested against the explicitly annotated PI-RADS values.

Multiple experiments were performed by changing various generative LLM models and using differing discriminative NER models for a broad comparative approach. Each pipeline extracted the maximum PI-RADS value and was tested against the explicitly annotated PI-RADS values. The best performing model types for differing architectures is shown in Table 1. The full evaluation output of all experiments performed are found in the supplemental materials (Supplementary Table 4). In addition, processing time and evaluation metrics were compared across pipeline configurations (Supplementary Table 5).

**Table 1:** Maximum PI-RADS evaluation metrics for 10 pipeline configurations.

| Experiment | Model | Pipeline Configuration | Macro Avg. |  |  | Avg. | Avg. |
| --- | --- | --- | --- | --- | --- | --- | --- |
|  |  |  | Precision | Recall | F1 | Secs per Text | Accuracy |
| 1 | Qwen3-14B | 2 | 0.962 | 0.964 | 0.962 | 34.286 | 0.955 |
| 2 | Qwen3-14B | 3 | 0.935 | 0.964 | 0.947 | 2.176 | 0.950 |
| 3 | Gemma 3n-E2B | 3 | 0.928 | 0.941 | 0.931 | 1.160 | 0.916 |
| 4 | Qwen3-8B | 3 | 0.888 | 0.948 | 0.910 | 1.094 | 0.923 |
| 5 | Qwen3-8B | 2 | 0.887 | 0.906 | 0.893 | 43.312 | 0.892 |
| 6 | Gemma 3n-E4B | 2 | 0.899 | 0.862 | 0.875 | 4.149 | 0.864 |
| 7 | spaCy (en_web_core_lg) | 1 | 0.907 | 0.846 | 0.869 | 0.134 | 0.873 |
| 8 | Transformers (en_web_core_trf) | 1 | 0.876 | 0.819 | 0.841 | 0.131 | 0.860 |
| 9 | Gemma 3n-E2B | 2 | 0.809 | 0.901 | 0.816 | 4.049 | 0.843 |
| 10 | Gemma 3n-E4B | 3 | 0.756 | 0.793 | 0.766 | 2.057 | 0.883 |

Performance characteristics across the ten pipeline configurations varied with average accuracies ranging from 84.3% to 95.5% accuracy in explicit PI-RADS outputs. Processing speed varied significantly across model configurations with traditional NER approaches demonstrating faster processing times (50-150 ms per text) compared to LLM-based methods (1-70 secs per text), suggesting LLM-based methods provide enhanced accuracy at the cost of computational resources.

## Discussion

This study provides one of the first head-to-head evaluations of three fundamentally different NLP paradigms—traditional NER, generative LLM extraction, and direct LLM inference—applied to the same clinical task of PI-RADS information extraction. The systematic evaluation of discriminative versus generative NLP pipelines reveals important insights for clinical deployment. First, discriminative NER models are significantly faster with modest reduction in accuracy (88% compared to 95%). Although generative LLMs slightly outperform NER methods in F1 score for extracting maximum PI-RADS values, the margin is small (≈4% difference), while NER models are 50–255× faster. For example, spaCy and Transformers-based NER pipelines process reports in 130 ms per document, compared to 1–70 seconds for LLMs. Speed is a critical determinant for real-world deployment in EHR environments and population-scale screening tasks. In large hospital systems, processing millions of radiology reports could become costly and require significant compute time such as days or months to run across a full radiology report document set. A potential trade-off could be using NER pipelines which enable scalable, low-resource extraction while maintaining clinically acceptable accuracy. Second, generative LLM pipelines deliver the highest accuracy but require more computing resources. Top-performing LLM configurations achieved F1 scores up to 96.2% and accuracy of 95.5%, placing them at the top of the performance rankings across 10 pipelines. This demonstrates that modern LLMs can reliably handle structured clinical inference tasks (e.g., identifying maximum PI-RADS) that require reasoning across heterogeneous free-text unstructured narratives. The choice of pipeline configuration depends on specific deployment requirements, including accuracy thresholds, processing speed constraints, and computational resource availability. Organizations with limited computational resources may benefit from discriminative-heavy configurations, while those prioritizing maximum accuracy may prefer generative approaches.

The current study builds on the findings of previous studies. Culnan et al^5^ developed a rule-based NLP algorithm to extract Gleason score, PI-RADS, PSA density, prostate volume, prostate dimensions, and biopsy core data from VA MRI and biopsy reports, achieving high F1 for PI-RADS (93.7), PSA density (99.5), prostate volume (95.7), and prostate dimensions (93.2). Our study, however, directly compared several discriminative (NER) vs generative (LLM) pipelines for the same MRI data elements, instead of a rule-based algorithm. With our findings, we were able to quantify speed vs accuracy trade-offs across 10 different configurations. Collado-Montanez et al^6^ created an automated text-based classification of malignancy scores in prostate MRI reports based on PI-RADS criteria, comparing traditional ML (XGBoost) vs. transformer based RoBERTa for NER. Our work built on this experience by comparing three pipeline paradigms, with NER (Transformers & spaCy), LLM extraction and LLM inference, showing structured information extraction and reasoning to identify maximum PIRADS value with PSA, prostate volume, and PSA density from unstructured reports.

Our study does have some limitations. This study focuses on a single clinical domain (prostate cancer screening) and specific dataset characteristics. This study does not examine temporal reasoning or tracking of PI-RADS changes over time, or multi-label extraction of narrative findings. Generalization to other clinical applications requires additional validation. We hope to utilize the current findings to investigate downstream clinical decision impact and cancer detection in future studies. Computational resource requirements for LLMs may present deployment challenges in resource-constrained clinical environments. Cost-benefit analysis should consider both improvements in accuracy and infrastructure requirements. Future work should explore dynamic pipeline selection based on input characteristics and confidence thresholds, enabling adaptive processing strategies that optimize performance for specific use cases. Some future considerations and research would be to test out additional hypothesis with mixed LLM & NER pipelines using various confidence cutoffs, as well as considering pre-trained NER models from previous research to validate and compare performance such as BioClinicalBERT or BI-RADS Bert.

This comprehensive evaluation of NLP pipeline configurations for prostate cancer screening demonstrates the potential of NER and LLM. The systematic comparison across ten experimental configurations provides practical guidance for clinical deployment decisions. Our contributions include (1) a systematic comparison of ten different NLP pipeline configurations for prostate cancer screening, (2) performance evaluation across multiple state-of-the-art language models, (3) speed and accuracy, and (4) comparative analysis with existing literature in automated radiology report processing. Key findings include the effectiveness of targeted LLM integration for complex reasoning tasks while maintaining traditional NER approaches for entity extraction efficiency. The optimal configuration varies based on specific deployment requirements and resource constraints. Future research should explore adaptive pipeline selection and expand evaluation to additional clinical domains to validate the generalizability of these findings. Additionally, we hope to evaluate hybrid strategies such as combining NER+LLM that may offer a real-world balance between accuracy and efficiency.

## Conclusion

In this study, we systematically compared discriminative and generative NLP pipelines for automated extraction of PI-RADS scores from prostate mpMRI radiology reports using a large, national VA dataset. Our findings demonstrate that while generative LLMs achieve the highest accuracy, discriminative NER-based approaches offer substantially greater efficiency and scalability. These results underscore the importance of aligning NLP pipeline design with health system resource constraints and workflow requirements and support the use of self-hosted, privacy-preserving NLP solutions to enable population-scale prostate cancer screening and research applications.

## Data Availability

All data produced in the present study are available upon reasonable request to the authors

