## Supplemental Tables for "Comparative analysis of discriminative and generative natural language processing pipelines for automated prostate magnetic resonance imaging reports"

**Supplemental Materials**

| | NER Data | |
|---|---|---|
| **Entity Type** | **Train Entities Quantity** | **Test Entities Quantity** |
| **PI-RADS Score** | 967 | 241 |
| **Prostate Dimensions** | 576 | 148 |
| **Prostate Volume** | 570 | 147 |
| **PSA** | 479 | 110 |
| **PSA Density** | 288 | 68 |
| *Total Entities* | *2880* | *714* |

Supplementary Table 1: Annotated entities identified in train and test sets for NER

| | Pipeline Evaluation Data | | | | | |
|---|---|---|---|---|---|---|
| **PI-RADS** | **1** | **2** | **3** | **4** | **5** | **Unavailable** |
| **Counts** | 9 | 59 | 55 | 45 | 46 | 85 |

Supplementary Table 2. PI-RADS annotations

### Full Evaluation Metrics for Multiple Pipeline Configurations

Below are the full evaluation metrics for the pipeline configurations along with additional models and evaluation results. The pipeline configuration number is associated with the sections under "Models and Configurations". The additional pipelines have a period followed by an additional number which specifies a model name difference from the numbered pipeline demonstrated in the main text body. The "Unavailable" results are when no PI-RADS was explicitly mentioned in the text and was annotated as "Unavailable".

| Experiment Number | Model Name | Pipeline Configuration | PI-RADS 1 | | | PI-RADS 2 | | |
|---|---|---|---|---|---|---|---|---|
| | | | Precision | Recall | F1 | Precision | Recall | F1 |
| 0 | Qwen2.5-14B | 3 | 1.000 | 1.000 | 1.000 | 0.918 | 0.949 | 0.933 |
| 1 | Qwen3-14B | 2 | 1.000 | 1.000 | 1.000 | 0.917 | 0.965 | 0.940 |
| 1.1 | Gemma3-12B | 2 | 1.000 | 1.000 | 1.000 | 0.915 | 0.947 | 0.931 |
| 1.2 | Qwen3-4B | 3 | 1.000 | 1.000 | 1.000 | 0.917 | 0.932 | 0.924 |
| 1.3 | Qwen2.5-14B | 2 | 1.000 | 0.889 | 0.941 | 0.915 | 0.947 | 0.931 |
| 2 | Qwen3-14B | 3 | 0.818 | 1.000 | 0.900 | 0.892 | 0.983 | 0.935 |
| 2.2 | Llama3-70B Instruct | 3 | 0.750 | 1.000 | 0.857 | 0.906 | 0.983 | 0.943 |
| 3 | Gemma 3n-E2B | 3 | 1.000 | 1.000 | 1.000 | 0.879 | 0.983 | 0.928 |
| 4 | Qwen3-8B | 3 | 0.643 | 1.000 | 0.783 | 0.921 | 0.983 | 0.951 |
| 4.1 | Mistral-7B | 2 | 0.750 | 1.000 | 0.857 | 0.927 | 0.895 | 0.911 |
| 4.2 | Gemma3-12B | 3 | 0.900 | 1.000 | 0.947 | 0.929 | 0.881 | 0.904 |
| 5 | Qwen3-8B | 2 | 0.818 | 1.000 | 0.900 | 0.922 | 0.825 | 0.870 |
| 5.1 | Qwen2.5-7B | 2 | 1.000 | 0.667 | 0.800 | 0.870 | 0.825 | 0.847 |
| 6 | Gemma 3n-E4B | 2 | 0.889 | 0.889 | 0.889 | 0.918 | 0.789 | 0.849 |
| 7 | spaCy (en_web_core_lg) | 1 | 1.000 | 0.667 | 0.800 | 0.922 | 0.797 | 0.855 |
| 8 | Transformers (en_web_core_trf) | 1 | 0.833 | 0.556 | 0.667 | 0.920 | 0.780 | 0.844 |
| 8.1 | Qwen2.5-7B | 3 | 1.000 | 0.556 | 0.714 | 0.970 | 0.542 | 0.696 |
| 9 | Gemma 3n-E2B | 2 | 0.346 | 1.000 | 0.514 | 0.930 | 0.930 | 0.930 |
| 10 | Gemma 3n-E4B | 3 | 0.818 | 1.000 | 0.900 | 0.817 | 0.983 | 0.892 |
| 10.1 | Llama3.2-3B | 2 | 1.000 | 0.778 | 0.875 | 0.813 | 0.912 | 0.860 |
| 10.2 | Qwen3-4B | 2 | 0.286 | 0.889 | 0.432 | 0.897 | 0.614 | 0.729 |

| | | | | | | | | |
|---|---|---|---|---|---|---|---|---|
| 10.3 | Mistral-7B | 3 | 0.417 | 0.556 | 0.476 | 0.815 | 0.373 | 0.512 |
| 10.4 | Llama3.2-3B | 3 | 0.000 | 0.000 | 0.000 | 0.000 | 0.000 | 0.000 |

Supplementary Table 3. Additional Pipeline Configuration Results for PI-RADS 1 & 2

| **Experiment** | **PI-RADS 3** | | | **PI-RADS 4** | | | **PI-RADS 5** | | | **PI-RADS Unavailable** | | |
|---|---|---|---|---|---|---|---|---|---|---|---|---|
| **Number** | **Precision** | **Recall** | **F1** | **Precision** | **Recall** | **F1** | **Precision** | **Recall** | **F1** | **Precision** | **Recall** | **F1** |
| 0 | 0.981 | 0.945 | 0.963 | 0.977 | 0.956 | 0.966 | 0.958 | 1.000 | 0.979 | 0.940 | 0.929 | 0.935 |
| 1 | 0.979 | 0.902 | 0.939 | 0.976 | 0.976 | 0.976 | 0.935 | 1.000 | 0.966 | 0.963 | 0.940 | 0.952 |
| 1.1 | 0.979 | 0.902 | 0.939 | 0.952 | 0.952 | 0.952 | 0.956 | 1.000 | 0.977 | 0.940 | 0.940 | 0.940 |
| 1.2 | 0.962 | 0.927 | 0.944 | 0.977 | 0.956 | 0.966 | 0.957 | 0.978 | 0.968 | 0.919 | 0.929 | 0.924 |
| 1.3 | 0.980 | 0.961 | 0.970 | 1.000 | 0.929 | 0.963 | 0.977 | 1.000 | 0.989 | 0.907 | 0.929 | 0.918 |
| 2 | 0.981 | 0.964 | 0.972 | 1.000 | 0.956 | 0.977 | 0.958 | 1.000 | 0.979 | 0.962 | 0.882 | 0.920 |
| 2.2 | 0.981 | 0.927 | 0.953 | 0.935 | 0.956 | 0.945 | 0.958 | 1.000 | 0.979 | 0.974 | 0.882 | 0.926 |
| 3 | 0.964 | 0.964 | 0.964 | 0.778 | 0.933 | 0.848 | 0.978 | 0.978 | 0.978 | 0.971 | 0.788 | 0.870 |
| 4 | 0.898 | 0.964 | 0.930 | 0.956 | 0.956 | 0.956 | 0.939 | 1.000 | 0.968 | 0.971 | 0.788 | 0.870 |
| 4.1 | 0.941 | 0.941 | 0.941 | 0.972 | 0.833 | 0.897 | 0.955 | 0.977 | 0.966 | 0.852 | 0.893 | 0.872 |
| 4.2 | 0.941 | 0.873 | 0.906 | 0.824 | 0.933 | 0.875 | 0.793 | 1.000 | 0.885 | 0.973 | 0.835 | 0.899 |
| 5 | 0.918 | 0.882 | 0.900 | 1.000 | 0.881 | 0.937 | 0.769 | 0.930 | 0.842 | 0.895 | 0.917 | 0.906 |
| 5.1 | 0.979 | 0.922 | 0.949 | 0.974 | 0.881 | 0.925 | 0.955 | 0.977 | 0.966 | 0.792 | 0.905 | 0.844 |
| 6 | 0.951 | 0.765 | 0.848 | 0.946 | 0.833 | 0.886 | 0.953 | 0.953 | 0.953 | 0.738 | 0.940 | 0.827 |
| 7 | 0.882 | 0.818 | 0.849 | 0.932 | 0.911 | 0.921 | 0.918 | 0.978 | 0.947 | 0.786 | 0.906 | 0.842 |
| 8 | 0.860 | 0.782 | 0.819 | 0.933 | 0.933 | 0.933 | 0.957 | 0.957 | 0.957 | 0.755 | 0.906 | 0.824 |

| | | | | | | | | | | | | |
|---|---|---|---|---|---|---|---|---|---|---|---|---|
| 8.1 | 0.974 | 0.673 | 0.796 | 0.977 | 0.956 | 0.966 | 0.939 | 1.000 | 0.968 | 0.631 | 0.965 | 0.763 |
| 9 | 0.754 | 0.961 | 0.845 | 0.891 | 0.976 | 0.932 | 0.955 | 0.977 | 0.966 | 0.979 | 0.560 | 0.712 |
| 10 | 0.981 | 0.964 | 0.972 | 0.754 | 0.956 | 0.843 | 0.958 | 1.000 | 0.979 | 0.965 | 0.647 | 0.775 |
| 10.1 | 0.759 | 0.863 | 0.807 | 0.886 | 0.929 | 0.907 | 0.527 | 0.907 | 0.667 | 0.641 | 0.298 | 0.407 |
| 10.2 | 0.968 | 0.588 | 0.732 | 0.966 | 0.667 | 0.789 | 0.473 | 0.814 | 0.598 | 0.776 | 0.786 | 0.781 |
| 10.3 | 0.907 | 0.709 | 0.796 | 0.412 | 0.889 | 0.563 | 0.724 | 0.913 | 0.808 | 0.645 | 0.471 | 0.544 |
| 10.4 | 0.111 | 0.018 | 0.031 | 0.210 | 0.733 | 0.327 | 0.463 | 0.804 | 0.587 | 0.574 | 0.318 | 0.409 |

Supplementary Table 4. Additional Pipeline Configuration Results for PI-RADS 3, 4, 5 and Unavailable

### NER Token level metrics on hold-out data

Below are the label metrics per token for the trained NER model against the hold-out dataset. It demonstrates the model evaluation performance using standard machine learning metrics. The support is the representative sample statistics for each token used in the hold-out data, i.e. the amount of tokens with the entity label in question.

| **Token Label Metrics** | | | | |
|---|---|---|---|---|
| **Label** | **precision** | **recall** | **f1-score** | **support** |
| None | 0.999 | 0.998 | 0.998 | 61772 |
| PI-RADS | 0.860 | 0.901 | 0.88 | 233 |
| PROSTATE_DIMENSIONS | 0.947 | 0.971 | 0.959 | 818 |
| PROSTATE_VOLUME | 0.887 | 0.916 | 0.901 | 155 |
| PSA | 0.833 | 0.882 | 0.857 | 102 |
| PSA_DENSITY | 0.855 | 0.951 | 0.900 | 62 |

| | | | | |
|---|---|---|---|---|
| *accuracy* | 0.997 | 0.997 | 0.997 | 0.997 |
| *macro avg* | 0.897 | 0.936 | 0.916 | 63142 |
| *weighted avg* | 0.997 | 0.997 | 0.997 | 63142 |

Supplementary Table 5. NER Performance Evaluation per Entity Type

### Runtime metrics per document quantity

Below are some results for multiple experiments sorted by shortest to longest time per document.

As seen below, the NER model pipelines are the fastest by a factor of 4 to their closest LLM counterparts and a factor of 255 times faster than the best performing LLM. The NER, in general, is less accurate which is tied directly to its ability to extract relevant entities, in this case the PI-RADS value.

Therefore, the NER loses ~8% accuracy but can perform 100k document extractions in about 3.7 hours while the best performing LLM would require 39 days to perform the same task. A single document is the concatenated report and impressions text. All tests were run on a Macbook Pro M3 with 48GB of RAM except for experiment 2.2.

| **Experiment Number** | **Model Name** | **Pipeline Configuration** | **Average Seconds Per Document** | **Average Days Per 100k Documents** |
|---|---|---|---|---|
| 8 | Transformers (en_web_core_trf) | 1 | 0.131 | 0.151 |
| 7 | spaCy (en_web_core_lg) | 1 | 0.134 | 0.155 |
| 10.4 | Llama3.2-3B | 3 | 0.494 | 0.572 |
| 8.1 | Qwen2.5-7B | 3 | 0.940 | 1.088 |

| | | | | |
|---|---|---|---|---|
| 4 | Qwen3-8B | 3 | 1.094 | 1.266 |
| 10.1 | Llama3.2-3B | 2 | 1.100 | 1.273 |
| 3 | Gemma 3n-E2B | 3 | 1.160 | 1.342 |
| 2.2 | Llama3-70B Instruct | 3 | 1.185 | 1.372 |
| 10.3 | Mistral-7B | 3 | 1.251 | 1.448 |
| 5.1 | Qwen2.5-7B | 2 | 1.669 | 1.932 |
| 0 | Qwen2.5-14B | 3 | 1.846 | 2.137 |
| 10 | Gemma 3n-E4B | 3 | 2.057 | 2.380 |
| 2 | Qwen3-14B | 3 | 2.176 | 2.519 |
| 4.1 | Mistral-7B | 2 | 3.383 | 3.915 |
| 4.2 | Gemma3-12B | 3 | 3.438 | 3.979 |
| 1.3 | Qwen2.5-14B | 2 | 3.594 | 4.159 |
| 9 | Gemma 3n-E2B | 2 | 4.049 | 4.686 |
| 6 | Gemma 3n-E4B | 2 | 4.149 | 4.802 |
| 1.1 | Gemma3-12B | 2 | 6.608 | 7.649 |
| 1.2 | Qwen3-4B | 3 | 32.919 | 38.101 |
| 1 | Qwen3-14B | 2 | 34.286 | 39.683 |
| 5 | Qwen3-8B | 2 | 43.312 | 50.130 |
| 10.2 | Qwen3-4B | 2 | 95.227 | 110.217 |

Supplementary Table 6. Additional pipeline results with per text prediction times.

### LLM Prompts for extractions and classifications

The following prompts were used in the pipelines with the LLM components for extraction and classification of texts. The '{multi_shot_examples}' was filled with additional examples found in the text to increase performance. Then the '{query}' was filled with the concatenated report + impressions text.

#### LLM Prompt for Pipeline Configuration 2

The prompt used for entity extraction in pipeline configuration 2 is shown below. It aims at creating a similar type of NER model used in pipeline 1, but instead relies on named entity extraction using an LLM with a predictable output structure. The numerical list output structure used a "type:name" string for ease of parsing.

```
You are a Named Entity Recognition System for Medical Documents.
You extract relevant medical terms from radiology reports for prostate cancer.

The relevant entities you extract are:

* PIRADS: A structured reporting scheme for MRIs in the evaluation of suspected prostate cancer in treatment. Values are integers from 1-5.
* PSA: Prostate specific antigen (PSA) is used as a tumor marker for prostate cancer. Values are floats; [e.g. 2.5, 4.5]
* PSA Density: A calculated diagnosis of prostate cancer resulting in a value in the units ng/mL.
* Prostate Dimension: Dimension of the prostate as a three valued pair, Anteroposterior (AP), Transverse (TRV), and Craniocaudal (CC). Using in the format [e.g. A x B x C]
* Prostate Volume: A volume of the prostate dimension. Values are in cc/mL.

Extract all the entities from the medical text.
Show only the numerical list starting from 1. with one extraction per line.
Each line should be the format <ENTITY_TYPE>: <EXTRACTED_VALUE>
Each entity type should be capitals

Be as concise as possible, mention only the values and entity types.
Do not add any explanations or logic about how you concluded your answers.
Do not make a bracked list of numbers.
Do not mention any additional information.

If No entities in the above list are found, respond only with 'NONE'.
```

If the definition of an entity is present, such as PI-RADS categories scoring values, ignore them entirely.

{multi_shot_examples}

Q: {query}

*Supplementary Prompt 1. Pipeline 2 Prompt*

The prompt below was used for maximum PI-RADS classification in pipeline configuration 3. It aims at extracting only PI-RADS scores for direct max detection. The numerical list output structure used was one PI-RADS per line.

```
You are a Medical Document Named Entity Recognition System.
You extract relevant medical terms from radiology reports.
Extract all PI-RADS scores from the text in a numerical list starting from 1.
Show only the numerical list starting from 1. with one extraction per line.
Be as concise as possible, mention only the numerical PIRADS values
Do not make a bracked list of numbers.
Do not mention any additional information.
If none are found, respond with 'NONE'.
If definitions of PI-RADS are present, ignore them entirely.

{multi_shot_examples}

Q: {query}
```

*Supplementary Prompt 2. Pipeline 3 Prompt*

| **Experiment** | **Model** | **100k Time** | **1m Time** |
|---|---|---|---|
| 8 | Transformers (en_web_core_trf) | 9 Days | 90 Days |
| 10.2 | Qwen3-4B | 18 Years | 181 Years |

*Supplementary Table 7: Comparison of Time to assess reports using different models*
